# Protocol and Statistical Analysis Plan for the ECMO-Free Trial: A Multicenter Randomized Controlled Trial

**DOI:** 10.64898/2025.12.01.25341410

**Authors:** Whitney D. Gannon, Ricardo Teijeiro-Paradis, Matthew E. Prekker, Melissa A. Vogelsong, Gary S. Schwartz, Mazen F. Odish, Nils P. Nickel, Sarah L. Bloom, Sydney J. Hansen, Vikram Fielding-Singh, Britton Blough, Robert L. Owens, Ramon Valles, Wren S. Adkisson, Bret D. Alvis, Matthew Bacchetta, Daniel J. Ford, Sean C. Gaudio, Christina A. Jelly, Janna S. Landsperger, Kaitlyn Lingle, Christy C. Noblit, Todd W. Rice, Lorenzo Del Sorbo, John W. Stokes, Joanna L. Stollings, Grace Van Winkle, Li Wang, Brant Imhoff, Matthew S. Shotwell, Eddy Fan, Matthew W. Semler, Jonathan D. Casey, ECMO-Free Investigators and the Pragmatic Critical Care Research Group

**Author notes:** Corresponding Author: Whitney D. Gannon, MSN MS, Division of Allergy, Pulmonary, and Critical Care Medicine Vanderbilt University Medical Center, Address: Vanderbilt University Medical Center C-1215 Medical Center North, Nashville, TN 37232. Collaborators for the ECMO-Free Trial Investigators and the Pragmatic Critical Care Research Group are listed in Section 1 of e-Appendix 1.

## Abstract

**Take-Home Points:** *Study Question:* Does a daily protocolized assessment of readiness for decannulation from venovenous extracorporeal membrane oxygenation (V-V ECMO) shorten the time to decannulation?

*Results:* This manuscript describes the protocol and statistical analysis plan for the ECMO-Free Trial, a multicenter randomized trial comparing a protocolized assessment of readiness to decannulate from ECMO versus usual care.

*Interpretation:* Specifying the protocol and full statistical analysis plan before completion of enrollment increases rigor, reproducibility, and transparency of trial results.

**Background:** Decannulation from venovenous extracorporeal membrane oxygenation (V-V ECMO) at the earliest, safe opportunity would be expected to improve patient outcomes. Currently, the manner in which clinicians liberate patients from V-V ECMO is highly variable. Whether performing a daily protocolized assessment of readiness for decannulation decreases the time to decannulation for patients on V-V ECMO is unknown.

**Research Question:** Does the performance of a daily protocolized assessment of readiness to decannulate from ECMO (an “ECMO-free protocol”) decrease the time to successful decannulation compared to usual care among patients receiving V-V ECMO?

**Study Design and Methods:** The ECMO-Free Trial is a pragmatic, multicenter, unblinded, parallel-group, randomized trial being conducted in 7 sites in North America. The trial compares the daily performance of an ECMO-free protocol in which patients who meet safety criteria have their sweep gas flow rate turned to 0 liters per minute (simulating the patient being “off ECMO”) to usual care among 225 adults receiving V-V ECMO. The primary outcome is the time to successful decannulation from ECMO within 60 days. The secondary outcome is the number of ECMO-free days to day 60. Enrollment began on September 7, 2022, and is expected to conclude in 2026.

**Interpretation:** The ECMO-Free Trial will provide important data on the effects of a protocolized assessment of readiness to decannulate vs usual care on the time to decannulation and other outcomes among patients receiving V-V ECMO. Specifying the protocol and statistical analysis plan before the conclusion of enrollment increases the rigor, reproducibility, and transparency of the trial.

**Trial Registry:** ClinicalTrials.gov; No. NCT05486559; URL: www.clinicaltrials.gov

## INTRODUCTION

The use of venovenous extracorporeal membrane oxygenation (V-V ECMO) for adults with severe respiratory failure has dramatically increased over the last decade. In 2024, more than 20,000 patients received ECMO across nearly 600 centers worldwide^1^. Advances in technology, increasing clinical experience, and data from randomized trials have improved the process by which patients are selected for V-V ECMO and the manner in which V-V ECMO is delivered^2,3^. In contrast, the process by which patients are assessed for readiness to decannulate from V-V ECMO represents an important knowledge gap.

Each additional day a patient receives V-V ECMO exposes the patient to an increased risk of bleeding, clotting, and infection^2,4^. Decannulation from ECMO at the earliest safe timepoint would be expected to improve clinical outcomes, reduce costs, and optimize resource allocation. Yet, limited data are available to inform how clinicians should approach liberating patients from V-V ECMO. As a result, the approaches to liberation from V-V ECMO vary significantly between clinicians, hospitals, and regions^5–7^.

In current clinical care, clinicians frequently approach liberation from V-V ECMO by identifying signs of lung recovery and initiating incremental reductions in blood flow rate, fraction of delivered oxygen, and sweep gas flow rate (referred to as “weaning”)^5–8^. This approach is described in guidelines from the Extracorporeal Life Support Organization, in expert opinion, and in small descriptive studies^5–7^. However, prior data suggest that clinicians underestimate patients’ readiness for liberation from organ support.^9,10^ For critical care therapies like sedation and invasive mechanical ventilation, incremental reductions in support (weaning) have been found to be inferior to protocolized daily assessments of readiness for discontinuation.^9–13^ Compared to incremental weaning, “spontaneous awakening trials” and “spontaneous breathing trials” dramatically shorten the duration of support, reduce intensive care costs, and improve patient outcomes^9–15^.

Recent studies have applied a daily assessment of readiness to decannulate patients from V-V ECMO. Two single-arm feasibility studies^16,17^ and two retrospective observational studies^18,19^ suggest that this approach may be feasible and safe and may identify candidates for decannulation earlier than occurs in usual care^16–19^. However, these uncontrolled studies are insufficient to determine whether a daily protocolized assessment of readiness to decannulate from V-V ECMO improves patient outcomes.

To address this knowledge gap, we designed the ECMO-Free Trial, which compares a daily protocolized assessment of readiness to decannulate from ECMO (ECMO-free protocol) to a liberation strategy directed by the clinical team (usual care) among 225 adults receiving V-V ECMO. We hypothesize that use of the ECMO-free protocol will decrease the time to successful decannulation compared to usual care.

## STUDY DESIGN AND METHODS

This article was written in accordance with Standard Protocol Items: Recommendations for Interventional Trials (SPIRIT) guidelines (Table 1 and Section 2 of e-Appendix 1).

**Table 1:** Schedule of Enrollment, Interventions, and Assessments in the ECMO-Free Trial.

|  | **Eligibility Screen** | **Randomization and Allocation** | **Daily Assessments** |  | **Decannulation and Post Decannulation** |  |  | **Final Outcome Assessment** |
| --- | --- | --- | --- | --- | --- | --- | --- | --- |
| Timepoint | Patient on V-V ECMO | Within 48 hours of V-V ECMO cannulation | Safety Screen | ECMO-Free Protocol | Decannulation | 24 hours after decannulation | 48 hours after decannulation | 60 days |
| Enrollment: |  |  |  |  |  |  |  |  |
| Eligibility screen | **X** |  |  |  |  |  |  |  |
| Consent |  | **X** |  |  |  |  |  |  |
| Enrollment |  | **X** |  |  |  |  |  |  |
| Allocation |  | **X** |  |  |  |  |  |  |
| Interventions: |  |  |  |  |  |  |  |  |
| ECMO-free protocol |  |  | **X** | **X** |  |  |  |  |
| Usual care |  |  | **X** |  |  |  |  |  |
| Assessments: |  |  |  |  |  |  |  |  |
| Baseline variables | **X** | **X** |  |  |  |  |  |  |
| On-study variables |  | **X** | **X** | **X** | **X** |  |  |  |
| Clinical outcomes |  |  |  |  | **X** | **X** | **X** | **X** |
| Adverse events |  | **X** | **X** | **X** | **X** | **X** | **X** | **X** |

### Study Design

The ECMO-Free Trial is a pragmatic, multicenter, unblinded, parallel-group randomized trial comparing a daily protocolized assessment of readiness to liberate from ECMO (ECMO-free protocol) versus usual care in 225 adults receiving V-V ECMO in 7 sites in North America. The trial is being conducted by the Pragmatic Critical Care Research Group (www.pragmaticcriticalcare.org). The trial was registered prior to initiation of enrollment (ClinicalTrials.gov Identifier: NCT05486559).

### Ethics and Regulatory Approval

The ECMO-Free Trial protocol was approved by the institutional review board (IRB) at Vanderbilt University Medical Center (IRB number: 220733) and by the IRB at each participating site.

For all patients, written informed consent is obtained from the patient or a surrogate decision maker prior to enrollment.

### Study Population

Critically ill adults receiving V-V ECMO in a participating intensive care unit are potentially eligible. The trial excludes patients who are pregnant, prisoners, less than 18 years old, or receiving ECMO as a bridge to transplant. A complete list of inclusion and exclusion criteria are provided in Table 2.

**Table 2:** Inclusion and Exclusion Criteria.

|  |
| --- |
| **Inclusion Criteria** |
| Patient is receiving venovenous extracorporeal membrane oxygenation |
| Patient is located in a participating unit of an adult hospital |
| **Exclusion Criteria** |
| Patient is known to be pregnant |
| Patient is known to be a prisoner |
| Patient is known to be less than 18 years old |
| Patient is receiving extracorporeal membrane oxygenation as a bridge to transplant |
| Patient is receiving a hybrid configuration that includes an arterial cannula |
| Patient has received venovenous extracorporeal membrane oxygenation for greater than 48 hours |

### Randomization and Treatment Allocation

Patients are randomized in a 1:1 ratio to the ECMO-free protocol group or the usual care group using a cloud-based randomization platform (REDCap) in permutated blocks of variable size, stratified by trial site. The study group assignment remains concealed to study personnel and treatment teams until after written informed consent has been obtained. Following randomization, the treatment team is notified of a patient’s group assignment. Given the nature of the intervention, patients, clinicians, and research personnel are aware of trial group assignment after randomization (unblinded).

## STUDY INTERVENTIONS

### ECMO-Free Protocol group

For patients in the ECMO-free protocol group, key study personnel perform a protocolized assessment of readiness for decannulation from V-V ECMO once daily. The ECMO-free protocol is a 3-step process of assessing readiness for decannulation from V-V ECMO: a safety screen, optimization of the non-ECMO respiratory support, and a trial off ECMO support (Figure 1).

**Figure 1.**
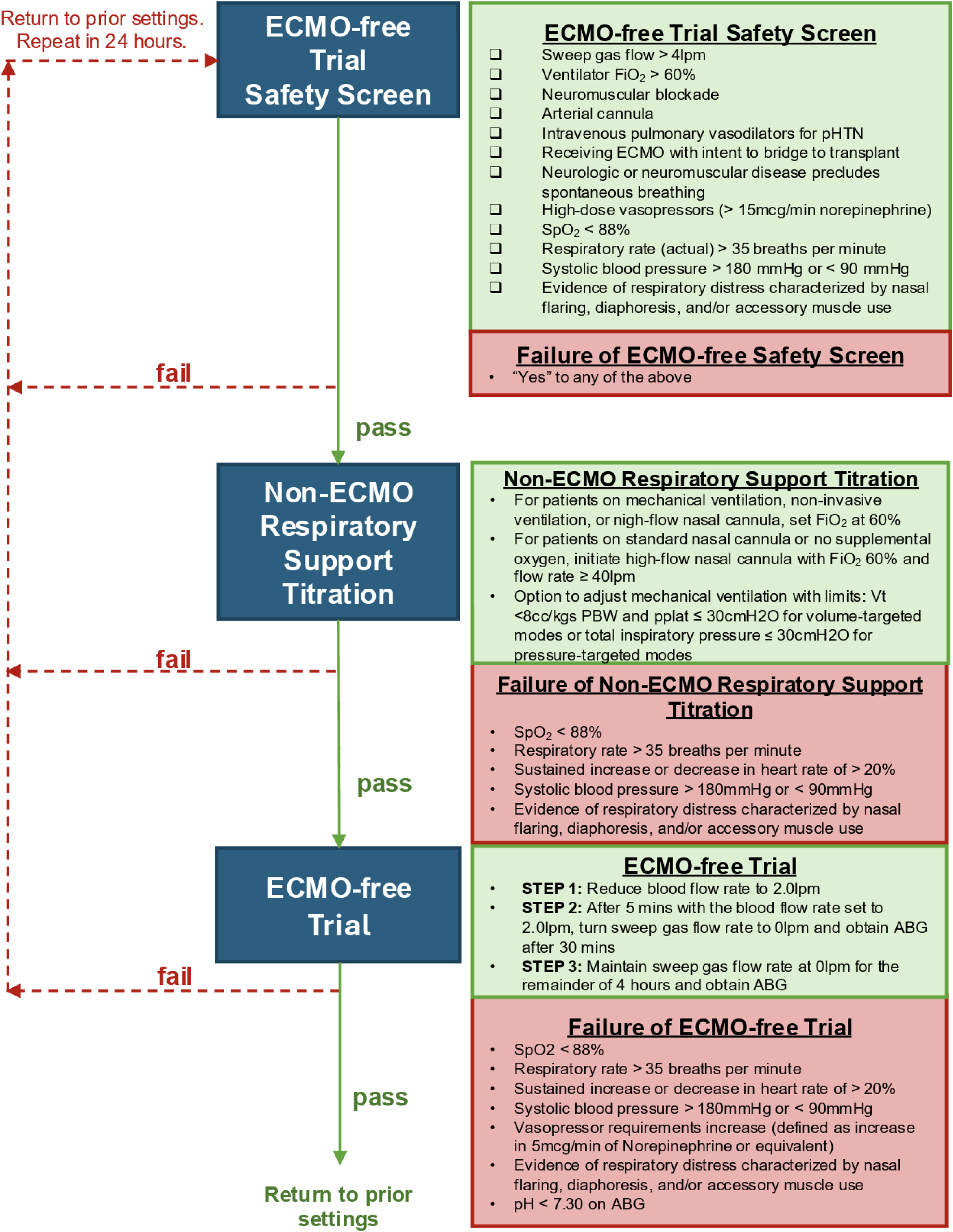
Flow chart showing the simplified beside tool used to apply the ECMO-free protocol during the trial.

To trial patients off ECMO support, the blood flow rate is first reduced to 2 L/min for five minutes (if applicable), after which the sweep gas flow is set to 0 L/min. When the sweep gas flow rate is turned to 0 L/min there is no gas exchange occurring through the membrane oxygenator, which simulates the patient being “off-ECMO”. The reduction of blood flow is an intermediary safety step that is performed to avoid the occurrence of rapid, severe hypoxemia observed among patients receiving high blood flow rates in a prior feasibility study^16^. Patients remain off ECMO support until the first of four hours or failure of the ECMO-free trial (defined using prespecified failure criteria; Section 3 e-Appendix 1). At the end of the ECMO-free protocol, the key study personnel who supervised the ECMO-free protocol verbally communicates the results to the clinical team. For patients who fail the ECMO-free trial (e.g., experience hypoxemia), study personnel immediately return the ECMO support and ventilator settings to prior levels. If the patient passes the ECMO-free trial, the sweep gas flow is continued at 0 L/min and all further clinical care is at the discretion of treating clinicians, including whether and when to resume ECMO support (e.g., increasing sweep gas flow) or proceed to decannulation from ECMO.

### Usual Care group

For patients in the usual care group, key study personnel perform only the daily safety screen. The results of the safety screen are not conveyed to clinicians, and the approaches to weaning and assessing readiness to decannulate from ECMO are at the discretion of treating clinicians (Section 4 e-Appendix 1).

### Duration of the intervention

The daily safety screen (both groups) and ECMO-free protocol (intervention group) are continued from enrollment to the first of decannulation, death, or 60 days after enrollment.

### Co-Interventions

All other aspects of how ECMO is provided at participating sites are determined by treating clinicians and local institutional clinical protocols for patient selection, cannulation, configuration, mechanical ventilation, sedation, anticoagulation, and post-decannulation care.

### Data Collection

For all patients in the trial, key study personnel review the electronic health record to collect data on baseline characteristics, the results of daily safety screens, and clinical outcomes, including date of decannulation, date of liberation from mechanical ventilation, date of ICU discharge, date of hospital discharge, and vital status. For patients assigned to the ECMO-free protocol group, study personnel not involved in the clinical care of the patient collect ECMO settings, respiratory support device settings, hemodynamics, and arterial blood gases during the performance of the ECMO-free protocol. All data are entered directly into the secure on-line REDCap database.

### Primary Outcome

The primary outcome is the time to successful decannulation from ECMO within 60 days, defined as the time from randomization (time zero) until the time of the final successful decannulation from ECMO within 60 days after enrollment. Decannulation from ECMO is classified as successful if a patient undergoes decannulation between randomization and 60 days and survives without ECMO until day 60. For patients who are decannulated and subsequently experience recannulation and subsequent decannulation, the time of successful decannulation from ECMO is based on the final decannulation. Ascertainment of data will cease at the time of hospital discharge. Data on the primary outcome will be censored at day 60.

The values for the primary outcome for patients who experience successful decannulation within 60 days range from 0.0 days to 60.0 days, with a smaller number of days considered to represent a better outcome than a larger number of days. For all patients who experience successful decannulation between randomization and 60 days and do not die between randomization and 60 days, the value for the time to decannulation will be calculated as the date and time (in hours and minutes) of decannulation minus the date and time (in hours and minutes) of randomization. All patients who remain on ECMO at 60 days or who die between randomization and 60 days (before or after decannulation from ECMO) will be treated as censored at day 60 for the primary outcome, such that in the analysis of the time to event, these patients will be considered to have experienced the worst possible outcome.

### Secondary Outcome

The secondary outcome is ECMO-free days to day 60, as defined by the number of days alive and free of ECMO to day 60 (Table 3). ECMO-free days will be calculated as 60 minus the number of calendar days from randomization to final decannulation. Patients receiving ECMO at day 60 will receive a value of “0”. Patients who die before day 60 or hospital discharge will receive a value of “0”.

**Table 3:** Study Outcomes.

|  |  |
| --- | --- |
| **Primary Outcome** | Time to successful decannulation from ECMO within 60 days, defined as time (in days) from randomization (time zero) until the time of the final successful decannulation from ECMO. Decannulation from ECMO will be considered to be successful if a patient undergoes decannulation and survives without ECMO until day 60. For patients who are decannulated and subsequently experience recannulation, the time of successful decannulation from ECMO will be based on the final decannulation. Data collection will cease at the time of hospital discharge or day 60. |
| **Secondary Outcome** | ECMO-free days to day 60, defined as 60 minus the number of days from randomization to final decannulation with patients who die before the first of day 60 or hospital discharge receiving "0" ECMO-free days |
| **Safety Outcomes** | Unsafe decannulation from ECMO, defined according to a previously published definition<sup>22</sup>. A patient will be classified as having experienced unsafe decannulation from ECMO if they experience any of the following within 48 hours of decannulation: V-V ECMO recannulation, sustained (> 4 hours) escalation of mechanical ventilation (change from a partially assisted mode to controlled mechanical ventilation, or dynamic driving pressure greater than or equal to 16 and delta change from previous setting of greater than or equal to 5 cm H<sub>2</sub>O, or increase in fraction of inspired oxygen to > 80%), use of rescue therapies (i.e., new need for paralysis and deep sedation, or inhaled pulmonary vasodilators, or high frequency oscillatory ventilation, or new/worsening hemodynamics requiring addition of any vasoactive agents with no evidence of sepsis or hypovolemia).<br>Recannulation within 30 days of decannulation |
| **Exploratory Outcomes** | Survival to decannulation<br>Respiratory support (ventilator and ECMO)-free days to day 60<br>ICU-free days to day 60<br>Ventilator-free days to day 60<br>Hospital-free days to day 60<br>60-day in-hospital mortality |

*Definitions of abbreviations:* ECMO = extracorporeal membrane oxygenation; ICU = intensive care unit

### Safety Outcomes

Trial protocol specifies two safety outcomes. The first is recannulation within 30 days of decannulation. The second is unsafe decannulation from ECMO according to a previously published definition^19^. A patient is classified as having experienced unsafe decannulation ECMO if they experience any of the following within 48 hours of decannulation: ECMO recannulation; sustained escalation of mechanical ventilation; use of rescue therapies; or new or worsening hemodynamics requiring addition of any vasoactive agents with no evidence of sepsis or hypovolemia (Table 3).

### Exploratory Outcomes

Table 3 reports the exploratory outcomes. Definitions of free-day outcomes are included in Section 5 of e-Appendix 1.

### Data and Safety Monitoring Board

The composition and responsibilities of the Data and Safety Monitoring Board (DSMB) are described in Section 6 of e-Appendix 1. The DSMB developed a charter, approved the trial protocol, and approved the trial monitoring plan. The DSMB has the authority to recommend the trial stop at any point, request additional data, request interim analyses, or request modifications to the study protocol. No interim analyses for efficacy or futility are planned. Additional details are provided in Section 6 of e-Appendix 1.

### Trial Stages

The ECMO-Free Trial was initially designed as a pilot trial at 3 sites to demonstrate the feasibility of conducting a multicenter randomized trial evaluating the previously developed ECMO-free protocol.^16^ Enrollment in the trial began on September 7, 2022. The pilot trial planned to enroll until the first of either (i) 60 patients enrolled or (ii) 12 months of enrollment at each of the 3 sites. Because the initiation of enrollment at one of the three sites was delayed, on November 16, 2023 the sample size was increased to 90 patients to allow sufficient time to evaluate enrollment at each of the 3 sites for at least 12 months. During the pilot phase, enrollment at each site exceeded the planned metrics, and it was determined that conducting a multicenter trial was feasible.

On November 11, 2024 the investigators revised the trial protocol from a three-center trial focused on feasibility to a definitive multicenter randomized trial evaluating the effect of the trial interventions on clinical outcomes. At the time of this transition, (i) the primary outcome was changed from the number of ECMO-free days to day 60 to the time to successful decannulation from ECMO within 60 days based on newly available statistical simulation from pre-trial data indicating that this outcome provided greater statistical power, (ii) the sample size was increased from 90 patients to 225 patients to provide sufficient statistical power to evaluate the effect of the ECMO-free protocol on the primary outcome (details in the section on Sample Size below), (iii) a Data Safety Monitoring Board was composed to monitor the conduct of the definitive multicenter trial, (iv) the enrollment window was changed from 24 to 48 hours following cannulation, and (iv) the number of enrolling sites was increased from 3 to 7. Neither the investigators nor the DSMB reviewed any analysis of trial data as part of the transition from a pilot trial to a definitive multicenter trial. Transitioning from a pilot feasibility phase to a definitive trial while including data from patients enrolled in the pilot phase is a well-accepted design known as an “internal pilot” study^20^. Prior consensus papers have noted that an internal pilot is an appropriate design when the eligibility criteria, intervention, and research context are all unlikely to change between the pilot phase and definitive trial, all of which were met in the case of the ECMO-Free Trial.

### Ancillary Study

An ancillary study of serum biomarkers was performed in a subset of patients enrolled in the ECMO-Free Trial. Results of this ancillary study will be reported separately from the results of the ECMO-Free Trial.

### Sample Size Estimation

The final sample size for the ECMO-Free Trial was calculated prior to the transition of the complete multicenter stage of the trial (details of the sample size estimate for the feasibility stage of the trial are provided in Section 7 of e-Appendix 1). For the primary outcome of time to successful decannulation from ECMO, the minimal difference that would be clinically important enough to justify the routine use of the ECMO-free protocol in clinical care is unknown. In simulations, performed on data on time to successful decannulation from ECMO among previous patients in the study settings prior to initiation of the study using the subdistribution hazard method to account for death as a competing event, we calculated that a total sample size of 225 patients (112 or 113 per group) would provide 80% statistical power at a two-sided alpha level of 0.05 to detect a hazard ratio of 1.68 for successful ECMO weaning in the ECMO-free protocol group compared to the usual care group. A hazard ratio of 1.68 was equivalent to a difference between groups of 1.3 days in the time to successful decannulation from ECMO. Additional details of the trial’s sample size estimates are in Section 7 of e-Appendix 1.

### Statistical Analysis Principles

Categorical variables will be presented as number and percentage. Continuous variables will be presented as mean ± standard deviation or median (interquartile range). We will analyze a single pre-specified primary outcome and a single pre-specified secondary outcome.

Consistent with recommendations of the Food and Drug Administration and the European Medicines Agency, each will be tested using a two-sided P value with a significance level of 0.05. For all other analyses except safety analyses, emphasis will be placed on the estimate of effect size with 95% confidence intervals, as recommended by the Internal Committee of Medical Journal Editors, and no corrections for multiple comparisons will be performed.

### Main Analysis of the Primary Outcome

The primary analysis will be an intention-to-treat comparison of patients randomized to the ECMO-free protocol group versus patients randomized to the usual care group with regard to the primary outcome of time to successful decannulation from ECMO with the use of an unadjusted Cox proportional hazards model. Patients who experience successful decannulation will be considered to have had an event, and the time of the event will be the date and time (in hours and minutes) of the final decannulation. Patients who remain on ECMO at 60 days or die between randomization and day 60 will be censored at day 60. Results will be presented with the use of a hazard ratio, 95% confidence intervals, and P value. Hazard ratio values greater than 1.0 will indicate earlier successful decannulation from ECMO for patients in the ECMO-free protocol group as compared with the usual care group (a better outcome with the ECMO-free protocol). A hazard ratio of 1.0 will indicate no difference in the primary outcome between the two trial groups. Hazard ratio values less than 1.0 will indicate later successful decannulation from ECMO in the ECMO-free protocol group versus the usual care group (a worse outcome with the ECMO-free protocol).

### Additional Analyses of the Primary Outcome

#### Adjusted analysis

We will repeat the main analysis of the primary outcome using a Cox proportional hazards model with adjustment for prespecified baseline covariates alone, trial site (strata) alone, and both baseline covariates and trial site (strata). Prespecified baseline covariates will include: age, sex, body mass index, acute indication for ECMO, presence of acute kidney injury prior to receipt of ECMO, simplified acute physiology score II at cannulation, Respiratory ECMO Survival Prediction Score at cannulation, pre-ECMO ventilator days, presence of septic shock, and ratio of the partial pressure of arterial oxygen to fraction of inspired oxygen prior to cannulation (Table 1).

#### Cumulative incidence analysis

We will perform a sensitivity analysis comparing patients randomized to the ECMO-free protocol group versus patients randomized to the usual care group with regard to the cumulative incidence of successful decannulation from ECMO and the cumulative incidence of death using the Fine and Gray competing risk model. The subdistribution hazard ratios will be estimated (with their 95% CIs) for both successful decannulation from ECMO and death.

### Effect Modification (Subgroup Analyses)

To evaluate whether pre-specified baseline variables modify the effect of trial group assignment on the primary outcome, we will repeat the main analysis of the primary outcome using a Cox proportional hazards model with the primary outcome as the dependent variable and independent variables of the trial group, the proposed effect modifier, and the interaction between the two. To account for non-linear relationships, continuous variables will be analyzed using restricted cubic splines with between 3 and 5 knots. Forest plots will be used to graphically display the analyses, and locally weighted regression or partial effects plots will be used to portray the association between continuous covariates and the outcome. In accordance with the Instrument for assessing the Credibility of Effect Modification Analyses (ICEMAN) recommendations^21^, we have prespecified the following variables as potential effect modifiers and hypothesized the direction of effect modification for each (Section 8 of e-Appendix 1):

1. Age (Continuous) – if dichotomized for presentation in forest plot then we will present as age greater or equal to 50 years versus age less than 50 years
2. Body mass index (weight [kg] / height [m]^2^) (Continuous) – if dichotomized for presentation in forest plot then we will present as body mass index greater or equal to 30 kg/m^2^ versus body mass index less than 30 kg/m^2^
3. Primary indication for ECMO (Acute respiratory distress syndrome / Post-lung transplant / Other)
4. Compliance at cannulation ([tidal volume/(plateau pressure-PEEP)] for patients receiving volume control; [tidal volume/driving pressure] for patients receiving pressure control) – if dichotomized for presentation in forest plot then we will present as compliance greater or equal to 20 mL/cm H_2_O versus compliance less than 20 mL/cm H_2_O
5. Partial pressure of arterial oxygen to fraction of inspired oxygen ratio (P:F) at cannulation (Continuous) – if dichotomized for presentation in forest plot then we will present as P:F greater or equal to 60 versus P:F 60
6. pH (Continuous) – if dichotomized for presentation in forest plot then we will present as pH greater or equal to 7.25 versus pH less than 7.25
7. Respiratory ECMO Survival Prediction (RESP) score at cannulation (Continuous) – if dichotomized for presentation in forest plot then we will present as RESP score greater or equal to 0 versus RESP score less than 0

### Analyses of Secondary, Safety, and Exploratory Outcomes

We will perform intention-to-treat comparisons of secondary, safety, and exploratory outcomes. Categorical outcomes will be compared using a chi-square test. Continuous outcomes will be compared with a Wilcoxon rank sum test.

### Handling of Missing Data

We anticipate that no data on the primary outcome will be missing. When data are missing for the secondary, safety, or exploratory outcomes, we will perform complete-case analysis, excluding cases where the data for the analyzed outcome are missing. There will be no imputation of missing outcome data. In adjusted analyses, missing data for covariates will be imputed using multiple imputation methods.

## DISCUSSION

We report the rationale, design, and analysis plan for the ECMO-Free Trial, a 225-patient randomized trial of a daily protocolized assessment of readiness to liberate from V-V ECMO vs usual care among critically ill adults receiving V-V ECMO. Important aspects of the design of the study population, study intervention, and selection of the comparator group warrant discussion.

The outcomes of V-V ECMO vary considerably based on the underlying cause of respiratory failure. For example, patients receiving V-V ECMO for ARDS have been reported to require ECMO for longer durations and experience higher rates of death than patients who receive V-V ECMO for asthma exacerbations or respiratory support following lung transplantation. Whether a protocolized assessment of readiness to liberate from V-V ECMO is more or less beneficial in patients with specific indications for V-V ECMO is unknown. Because no standardized approaches to weaning exist for any patient population receiving V-V ECMO and all patients who receive V-V ECMO may benefit from a protocolized assessment of readiness for decannulation, the ECMO-Free Trial has been designed with broad eligibility criteria that include all patients receiving V-V ECMO for bridge to recovery except those for whom interrupting the sweep gas flow would be unsafe (e.g., patients with an arterial ECMO cannula). The trial analysis includes prespecified subgroup analyses to evaluate whether specific patient characteristics, such as indication for ECMO, modify the effectiveness of the intervention.

The minimum duration of time a patient should spend off ECMO (with a sweep gas flow of 0 L/min) to establish readiness for decannulation is unknown. In this trial, we chose an ECMO-free trial duration of 4 hours based on an observational study suggesting that this duration was sufficient to assess eligibility for decannulation^22^. When a patient passes the ECMO-free trial, this information is conveyed to their clinical team, but the trial does not mandate decannulation. Decannulation and recannulation are both associated with significant risk of complication, and clinicians may have additional considerations (e.g., trajectory of illness, planned procedures, etc.) that preclude decannulation of a patient who passes an ECMO-free trial. The approach used in this trial, in which the intervention is limited to conducting the ECMO-free trial and communicating the results of the test to treating clinicians, is similar to the approach used in prior trials that established the clinical benefits of spontaneous awakening and breathing trials from mechanical ventilation^10,13^.

The manner in which ECMO is weaned and discontinued in routine clinical care varies within and between centers. Current approaches generally rely on clinicians to identify signs of lung recovery and initiate gradual reductions in the blood flow rate and sweep gas flow rate at the discretion of the treating clinicians.^5–7^ Because no standardized protocols for weaning from ECMO exist, the ECMO-free protocol is being compared in this trial to usual care. Allowing clinicians to use the same approach to weaning they would use in routine clinical care maximizes generalizability. However, it could lead to contamination if clinicians adopt aspects of the ECMO-free protocol in the usual care group. To minimize contamination, the ECMO-free protocol is delivered by study team members rather than the care team and engagement with the clinical team for patients randomized to the usual care group was limited. To evaluate for contamination the trial prospectively records, in both groups, the time spent each day with a sweep gas flow of 0 L/min in both groups.

The primary outcome of the ECMO-Free Trial is time to successful decannulation from ECMO within 60 days. This outcome was chosen because (i) it is important to patients and clinicians, (ii) has been used in prior trials of V-V ECMO^23^, (iii) has been recognized as an important outcome in an international, multidisciplinary, modified Delphi study focused on core outcome measures for ECMO research^24^, (iv) is closely linked to the trial intervention, and (v) simulations on prior patients in the study setting suggest that it provides superior statistical power to alternative outcomes such as number of days alive and free from ECMO. Additional analyses will evaluate for the effect of the competing risk of mortality on the primary outcome and of the intervention on secondary and exploratory outcomes.

### Interpretation

The ECMO-Free Trial will provide important insight regarding the effect of a daily protocolized assessment of readiness to decannulate from V-V ECMO vs usual care on time to decannulation and other clinical outcomes among critically ill adults receiving V-V ECMO. To improve the transparency and interpretation of trial results, this manuscript presenting the protocol and statistical analysis plan for the ECMO-Free Trial has been completed prior to the conclusion of patient enrollment.

## Data Availability

All data produced in the present study are available upon reasonable request to the authors.

