## Supplement File for "Protocol and Statistical Analysis Plan for the ECMO-Free Trial: A Multicenter Randomized Controlled Trial"

#### **1. LIST OF ECMO-FREE INVESTIGATORS**

Coordinating Center: *Clinical Coordinating Center (Vanderbilt University Medical Center, Nashville TN)* – Jonathan D. Casey, MD MSc* (Director, Coordinating Center); Matthew W. Semler, MD MSc* (Chair, Steering Committee); Todd W. Rice, MD MSc*; Grace Van Winkle, MPH*; Sydney Lagalante, MPH; Kaitlyn R. Whitaker, MS; Cheryl L. Gatto, PhD; Data Coordinating Center (Vanderbilt University Medical Center, Nashville TN) – Brant Imhoff, MS*; Li Wang, MS*; Matthew S. Shotwell, PhD*

Toronto General Hospital: Ricardo Teijeiro-Paradis, MD*; Eddy Fan, MD PhD*, Lorenzo Del Sorbo, MD*

Hennepin County Medical Center: Matthew E. Prekker, MD*; Sydney J. Hansen, MD*; Laurynn Giles, BA; Mary O’Rourke, BA; Heidi Erickson, MD; Kenneth Dodd, MD; Beth Heather, MSN, RN, CCRN; Audrey Hendrickson, MPH

Stanford University: Melissa A. Vogelsong, MD*; Vikram Fielding-Singh, MD, JD*; Brandon A. Guenthart, MD; Joseph Simmons, PA-C, Brenda Rocha, ACNP; Arely Campos-Melendez, BS

University of California, San Diego: Mazen Odish, MD*; Robert L. Owens, MD*; Travis Pollema, DO

Baylor, Scott, and White, Dallas: Gary Schwartz, MD*; Britton Blough, MD*; Kaitlyn Lingle, RN, CCRN-CSC*

Texas Tech University: Nils P Nickel, MD*, Ramon Valles, MD*

Vanderbilt University Medical Center: Whitney D. Gannon, MS, MSN*; Bret D. Alvis, MD, MSc*; Sarah Bloom*; Sean C. Gaudio, MD*; Christina A. Jelly, MD*; John W. Stokes, MD*; Matthew Bacchetta, MD, MBA*; Daniel J. Ford, AG-ACNP*; Janna Landsperger*; Wren S. Adkisson, AG-ACNP*; Joanna L. Stollings, PharmD*; Yatrik J. Patel, MD; Christy C. Noblit, ACNP-BC*; Sean Francois, MD; William D. Tucker, MD; Brandon Petree, DO; Enock Adjei, MD; Amir Dereshgi, MD

*Denotes members of the writing committee who are listed as authors on the manuscript, the remainder of the ECMO-Free Trial investigators represent collaborators

#### **2. SPIRIT 2025 CHECKLIST**

SPIRIT 2025 Checklist: Recommended items to address in a clinical trial protocol and related documents*

| **Section / Topic** | **No** | **SPIRIT 2025 checklist item description** | **Reported on page no.** |
| --- | --- | --- | --- |
| **Administrative information** | | |  |
| Title and structured summary | 1a | Title stating the trial design, population, and interventions, with identification as a protocol | 1 |
|  | 1b | Structured summary of trial design and methods, including items from the World Health Organization Trial Registration Data Set | 6-9 |
| Protocol version | 2 | Version date and identifier | NA |
| Roles and responsibilities | 3a | Names, affiliations, and roles of protocol contributors | 1, Section 1 e-Appendix 1 |
|  | 3b | Name and contact information for the trial sponsor | NA |
|  | 3c | Role of trial sponsor and funders in design, conduct, analysis, and reporting of trial; including any authority over these activities | 2 |
|  | 3d | Composition, roles, and responsibilities of the coordinating site, steering committee, endpoint adjudication committee, data management team, and other individuals or groups overseeing the trial, if applicable | 1, 2, Section 1 e-Appendix 1 |
| **Open science** | | |  |
| Trial registration | 4 | Name of trial registry, identifying number (with URL), and date of registration. If not yet registered, name of intended registry | 4, 6 |
| Protocol and statistical analysis plan | 5 | Where the trial protocol and statistical analysis plan can be accessed | 4, 6 |
| Data sharing | 6 | Where and how the individual de-identified participant data (including data dictionary), statistical code, and any other materials will be accessible | NA |
| Funding and conflicts of interest | 7a | Sources of funding and other support (e.g., supply of drugs) | 2 |
|  | 7b | Financial and other conflicts of interest for principal investigators and steering committee members | 2 |
| Dissemination policy | 8 | Plans to communicate trial results to participants, healthcare professionals, the public, and other relevant groups (e.g., reporting in trial registry, plain language summary, publication) | NA |
| **Introduction** | | |  |
| Background and rationale | 9a | Scientific background and rationale, including summary of relevant studies (published and unpublished) examining benefits and harms for each intervention) | 5-6 |
|  | 9b | Explanation for choice of comparator | 5-6, 8 |
| Objectives | 10 | Specific objectives related to benefits and harms | 6 |
| **Methods: Patient and public involvement, trial design** | | |  |
| Patient and public involvement | 11 | Details of, or plans for, patient or public involvement in the design, conduct, and reporting of the trial | NA |
| Trial design | 12 | Description of trial design including type of trial (e.g., parallel group, crossover), allocation ratio, and framework (e.g., superiority, equivalence, non-inferiority, exploratory) | 6 |
| **Methods: Participants, interventions, and outcomes** | | |  |
| Trial setting | 13 | Settings (e.g., community, hospital) and locations (e.g., countries, sites) where the trial will be conducted | 7 |
| Eligibility criteria | 14a | Eligibility criteria for participants | 7, Table 2 |
|  | 14b | If applicable, eligibility criteria for sites and for individuals who will deliver the interventions (e.g., surgeons, physiotherapists) | 7, 8 |
| Intervention and comparator | 15a | Intervention and comparator with sufficient details to allow replication including how, when, and by whom they will be administered. If relevant, where additional materials describing the intervention and comparator (e.g., intervention manual) can be accessed | 7, 8, Section 3, 4 e-Appendix 1 |
|  | 15b | Criteria for discontinuing or modifying allocated intervention/comparator for a trial participant (e.g., drug dose change in response to harms, participant request, or improving/worsening disease) | NA |
|  | 15c | Strategies to improve adherence to intervention/comparator protocols, if applicable, and any procedures for monitoring adherence (e.g., drug tablet return, sessions attended) | NA |
|  | 15d | Concomitant care that is permitted or prohibited during the trial | 9 |
| Outcomes | 16 | Primary and secondary outcomes, including the specific measurement variable (e.g., systolic blood pressure), analysis metric (e.g., change from baseline, final value, time to event), method of aggregation (e.g., median, proportion), and time point for each outcome | 9-11 |
| Harms | 17 | How harms are defined and will be assessed (e.g., systematically, non-systematically) | 10 |
| Participant timeline | 18 | Time schedule of enrollment, interventions (including any run-ins and washouts), assessments, and visits for participants. A schematic diagram is highly recommended (see Figure) | Figure 1 |
| Sample size | 19 | How sample size was determined, including all assumptions supporting the sample size calculation | 11-13 |
| Recruitment | 20 | Strategies for achieving adequate participant enrollment to reach target sample size | NA |
| **Methods: Assignment of interventions** | | |  |
| Randomization: |  |  |  |
| Sequence generation | 21a | Who will generate the random allocation sequence and the method used | 7 |
|  | 21b | Type of randomization (simple or restricted) and details of any factors for stratification. To reduce predictability of a random sequence, other details of any planned restriction (e.g., blocking) should be provided in a separate document that is unavailable to those who enroll participants or assign interventions | 7 |
| Allocation concealment  mechanism | 22 | Mechanism used to implement the random allocation sequence (e.g., central computer/telephone; sequentially numbered, opaque, sealed containers), describing any steps to conceal the sequence until interventions are assigned | 7 |
| Implementation | 23 | Whether the personnel who will enroll and those who will assign participants to the interventions will have access to the random allocation sequence | 7 |
| Blinding | 24a | Who will be blinded after assignment to interventions (e.g., participants, care providers, outcome assessors, data analysts) | 7 |
|  | 24b | If blinded, how blinding will be achieved and description of the similarity of interventions | NA |
|  | 24c | If blinded, circumstances under which unblinding is permissible, and procedure for revealing a participant’s allocated intervention during the trial | NA |
| **Methods: Data collection, management, and analysis** | | |  |
| Data collection methods | 25a | Plans for assessment and collection of trial data, including any related processes to promote data quality (e.g., duplicate measurements, training of assessors) and a description of trial instruments (e.g., questionnaires, laboratory tests) along with their reliability and validity, if known. Reference to where data collection forms can be accessed, if not in the protocol | 9 |
|  | 25b | Plans to promote participant retention and complete follow-up, including list of any outcome data to be collected for participants who discontinue or deviate from intervention protocols | 9 |
| Data management | 26 | Plans for data entry, coding, security, and storage, including any related processes to promote data quality (e.g., double data entry; range checks for data values). Reference to where details of data management procedures can be accessed, if not in the protocol | 9 |
| Statistical methods | 27a | Statistical methods used to compare groups for primary and secondary outcomes, including harms | 13-16 |
|  | 27b | Definition of who will be included in each analysis (e.g., all randomized participants), and in which group | 13-16 |
|  | 27c | How missing data will be handled in the analysis | 16 |
|  | 27d | Methods for any additional analyses (e.g., subgroup and sensitivity analyses) | 15, 16 Section 9. e-Appendix 1 |
| **Methods: Monitoring** | | |  |
| Data monitoring committee | 28a | Composition of data monitoring committee (DMC); summary of its role and reporting structure; statement of whether it is independent from the sponsor and funder; conflicts of interest and reference to where further details about its charter can be found, if not in the protocol. Alternatively, an explanation of why a DMC is not needed | 11, Section 6. e-Appendix 1 |
|  | 28b | Explanation of any interim analyses and stopping guidelines, including who will have access to these interim results and make the final decision to terminate the trial | NA |
| Trial monitoring | 29 | Frequency and procedures for monitoring trial conduct. If there is no monitoring, give explanation | NA |
| **Ethics** | | |  |
| Research ethics approval | 30 | Plans for seeking research ethics committee/institutional review board approval | 7 |
| Protocol amendments | 31 | Plans for communicating important protocol modifications to relevant parties | NA |
| Consent or assent | 32a | Who will obtain informed consent or assent from potential trial participants or authorized proxies, and how | 7 |
|  | 32b | Additional consent provisions for collection and use of participant data and biological specimens in ancillary studies, if applicable | NA |
| Confidentiality | 33 | How personal information about potential and enrolled participants will be collected, shared, and maintained in order to protect confidentiality before, during, and after the trial | Section 11 e-Appendix 1 |
| Ancillary and post-trial care | 34 | Provisions, if any, for ancillary and post-trial care, and for compensation to those who suffer harm from trial participation | NA |

### **SUPPLEMENTAL METHODS**

#### **3. Study Interventions for the ECMO-Free Protocol Group**

For patients assigned to the ECMO-free protocol group, trained study personnel perform the ECMO-free protocol daily. The ECMO-Free protocol is a 3-step process of assessing readiness for decannulation from V-V ECMO and detailed here:

Phase 1: ECMO-free Safety Screen:

Study personnel perform the ECMO-free Safety Screen daily from enrollment to decannulation at the same time each day (typically between 6:00 AM and 10:00 AM local time). If enrollment occurs after 10:00 AM local time, the first ECMO-free Safety Screen is performed the following calendar day. The ECMO-free Safety Screen was designed to (i) ensure patient safety, (ii) maximize opportunities for patients to proceed to the ECMO-free protocol and be evaluated for readiness for decannulation, and (iii) use variables that are readily available in the electronic health record so that the safety screen could be completed remotely (without in-person interaction with the patient or treating clinicians). Each day study personnel assess for the presence of any of the following safety screen failure criteria:

- Sweep gas flow rate greater than 4LPM
- Fraction of inspired oxygen greater than 60%
- Receipt of neuromuscular blockade
- Presence of an arterial ECMO cannula
- Receipt of intravenous pulmonary vasodilators for pulmonary hypertension
- Receipt of ECMO with the intent to bridge to transplant
- Presence of a neurologic or neuromuscular disease that precludes spontaneous breathing
- Receipt of high-dose vasopressors (greater than 15mcg/min of Norepinephrine or equivalent)
- Oxygen saturation less than 88%
- Respiratory rate greater than 35 breaths per minute
- Systolic blood pressure greater than 180mmHg or less than 90mmHg
- Evidence of respiratory distress characterized by nasal flaring, diaphoresis, and/or accessory muscle use.

Results of the ECMO-free Safety Screen are prospectively recorded by study personnel. Study personnel are trained to evaluate for safety criteria when the patient is at a “steady state” (ie, not during reversible periods of discomfort/instability related to physical therapy, turning or suctioning). If the patient meets one or more ECMO-free Safety Screen criteria, the patient is considered to have failed the ECMO-free Safety Screen on that day. In the absence of any ECMO-free Safety Screen criteria, the patient is considered to have passed the ECMO-free Safety Screen on that day and proceeds to Phase 2: Non-ECMO respiratory support titration.

Phase 2: Non-ECMO Respiratory Support Titration:

When a patient does not meet any Safety Screen failure criteria, they progress to Phase 2: Non-ECMO Respiratory Titration. The aim of Phase 2 is to optimize non-ECMO respiratory support to maximize the likelihood of passing Phase 3 (the ECMO-free trial) and demonstrating readiness for decannulation. During Phase 2, the fraction of inspired oxygen is increased to 0.60 (if set below 0.60). For patients receiving standard oxygen therapy, high flow nasal cannula is initiated at a flow rate of 40 liters per minute and an FiO2 of 0.60. For patients receiving ultra-lung protective ventilation, ventilator settings are increased to standard settings, targeting tidal volumes <8cc/kgs predicted body weight and plateau pressures ≤30cmH_2_O for patients receiving a volume-targeted mode and a total inspiratory pressure (driving pressure plus positive end-expiratory pressure) ≤30cmH_2_O for patients receiving a pressure-targeted mode. For patients already receiving standard mechanical ventilation settings, no titration is required. While the goal of Phase 2 is to increase support and increase the safety margin for Phase 3 (the ECMO-free trial), patients can develop clinical instability during Phase 2, which is considered to be evidence that it would be unsafe to progress to Phase 3. Failure criteria during Phase 2 include: oxygen saturation less than 88%, respiratory rate greater than 35 breaths per minute, sustained increase or decrease in heart rate of greater than 20%, systolic blood pressure greater than 180mmHg or less than 90mmHg, or evidence of respiratory distress characterized by nasal flaring, diaphoresis, and/or accessory muscle use. Presence or absence of failure criteria is recorded prospectively. If one or more criteria are present, the patient is considered to have failed Phase 2. In the absence of these criteria, the patient is considered to have passed Phase 2, and the patient proceeds to Phase 3: the ECMO-free trial.

Phase 3: The ECMO-free Trial:

Phase 3 of the intervention is the ECMO-free trial in which ECMO support is rapidly weaned, and the patient’s respiratory status is evaluated without any respiratory support from V-V ECMO. In Phase 3, the blood flow rate is weaned to 2LPM (unless the blood flow rate is already ≤2LPM). If weaning of the flow is required, the patient is monitored for five minutes before proceeding with weaning of the sweep gas flow. Next, the sweep gas flow is rapidly weaned to 0LPM (simulating the patient being “off ECMO”). If desired, once the sweep gas flow rate is 0LPM, the blood flow rate can be returned to prior settings (to avoid a theoretical increase in the risk of circuit thrombosis at lower blood flow rates). Patients are continually monitored while on 0LPM sweep gas flow for the presence of any failure criteria (listed below):

- Oxygen saturation less than 88%
- Respiratory rate greater than 35 breaths per minute
- Sustained increase or decrease in heart rate of greater than 20%
- Increase in vasopressor requirements (defined as greater than 5mcg/min of Norepinephrine or equivalent)
- pH less than 7.30 on an arterial or venous blood gas
- Evidence of respiratory distress characterized by nasal flaring, diaphoresis, and/or accessory muscle use

The patient’s clinical status during the ECMO-free trial and the results of the ECMO-free trial are recorded prospectively. Study personnel remain at the bedside for at least 30 minutes after the sweep gas flow is weaned to 0L/min. If the patient remains stable at 30 minutes, monitoring occurs according to local clinical protocols. The ECMO-free trial continues until the first of 4 hours or failure. An arterial or venous blood gas is obtained at 30 minutes and 4 hours. For patients who fail the ECMO-free trial (e.g., experience hypoxemia), study personnel immediately return the ECMO support and ventilator settings to prior levels. If the patient remains on the ECMO-Free trial for 4 hours without meeting failure criteria, the clinical team is notified that the patient has passed the ECMO-free trial. The sweep gas flow rate is continued at 0L/min and further clinical management is at the discretion of the clinical team, including whether and when to resume ECMO support or proceed to decannulation.

#### **4. Study Interventions for the Usual Care Group**

For patients in the Usual Care Group, study personnel perform only the daily safety screen. The results of the safety screen are not conveyed to clinicians, and the approaches to weaning and assessing readiness to decannulate from ECMO are at the discretion of treating clinicians.

#### **5. Definitions of Free-Day Outcomes**

*ECMO-free days to day 60 (EFDs):* ECMO-Free Days are defined as the number of calendar days, between enrollment and 60 days after enrollment, on which the patient is alive and free of extracorporeal membrane oxygenation (ECMO). If a patient is liberated from ECMO, returns to ECMO and subsequently is liberated from ECMO again prior to day 60, the number of EFDs will be counted from the end of the last period of ECMO to day 60. If the patient is receiving ECMO at day 60 or dies prior to day 60, EFDs are 0. If a patient is discharged while receiving ECMO, EFDs are 0. Outcome ascertainment ends at 60 days or hospital discharge, whichever occurs first.

*Ventilator-free days to day 60 (VFDs):* VFDs are defined as the number of calendar days, between enrollment and 60 days after enrollment, on which the patient is alive and free of invasive mechanical ventilation. If a patient is liberated from invasive mechanical ventilation, returns to invasive mechanical ventilation and subsequently is liberated from invasive mechanical ventilation again prior to day 60, the number of VFDs will be counted from the end of the last period of invasive mechanical ventilation to day 60. If the patient is receiving invasive mechanical ventilation at day 60 or dies prior to day 60, VFDs are 0. If a patient is discharged while receiving invasive mechanical ventilation, VFDs are 0. Outcome ascertainment ends at 60 days or hospital discharge, whichever occurs first.

*Respiratory support-free days to day 60 (RSFDs):* RSFDs are defined as the number of calendar days, between enrollment and 60 days after enrollment, on which the patient is alive and free of both invasive mechanical ventilation and ECMO. If a patient is liberated from both invasive mechanical ventilation and ECMO, returns to invasive mechanical ventilation and/or ECMO and subsequently is liberated from invasive mechanical ventilation and/or ECMO again prior to day 60, the number of RSFDs will be counted from the end of the last period of invasive mechanical ventilation and/or ECMO to day 60. If the patient is receiving invasive mechanical ventilation and/or ECMO at day 60 or dies prior to day 60, RSFDs are 0. If a patient is discharged while receiving invasive mechanical ventilation and/or ECMO, RSFDs are 0. Outcome ascertainment ends at 60 days or hospital discharge, whichever occurs first.

*ICU-free days to day 60 (ICU-FDs):* ICU-FDs are defined as the number of calendar days, between enrollment and 60 days after enrollment, on which the patient is alive and not admitted to an intensive care unit after the patient’s final transfer out of the intensive care unit. Patients who are never transferred out of the intensive care unit receive a value of 0. Patients who die before day 60 receive a value of 0. For patients who are transferred out of the ICU, return to an ICU, and are subsequently transferred out of the ICU again prior to day 60, ICU-free days are counted from the date of final transfer out of the ICU. Outcome ascertainment ends at 60 days or hospital discharge, whichever occurs first.

*Hospital-free days to day 60 (Hospital-FDs):* Hospital-FDs are defined as the number of calendar days, between enrollment and 60 days after enrollment, on which the patient is alive and not admitted to the hospital. Patients who are never transferred out of the hospital receive a value of 0. Patients who die before day 60 receive a value of 0. Outcome ascertainment ends at 60 days or hospital discharge, whichever occurs first.

#### **6. Composition and Responsibilities of the Data Safety Monitoring Board**

The Data and Safety Monitoring Board (DSMB) consists of members with expertise in pulmonary and critical care medicine, extracorporeal membrane oxygenation, biostatistics, and clinical trials. All members of the DSMB are voting members. The DSMB approved the trial protocol and approved the trial monitoring plan at the time of the transition from the feasibility stage to the definitive, multicenter stage of the ECMO-Free Trial. The DSMB has the ability at any point to recommend that the trial end, be modified, or continue unchanged.

The principal role of the DSMB is to assure the safety of patients in the trial. The DSMB monitors data from the trial at annual meetings, reviews and assesses the performance of its operations, and makes recommendations about:

1. Participant safety and risk/benefit ratio of study procedures and interventions
2. Initial approval of the protocol and subsequent amendments (with specific attention to study population, intervention, and study procedures)
3. Adherence to the protocol requirements
4. Completeness, quality, and planned analysis of data
5. Ancillary study burden on participants and main study
6. Possible early termination of the trial because of new external information, early attainment of study objectives, safety concerns, or inadequate performance

#### **7. Details of Sample Size Calculation**

Sample Size Estimate for the Feasibility Stage

The feasibility stage was initially planned to continue to the first of 60 patients or 12 months of enrollment at each study site. These metrics were estimated to be sufficient to demonstrate the feasibility of enrollment and intervention delivery in a multicenter trial of weaning from V-V ECMO. Because the initiation of enrollment was significantly delayed at one of the three sites, the number of patients permitted to be enrolled in the feasibility stage was increased from 60 to 90, to ensure that each vanguard site was able to enroll patients for one full year.

Transition to a Multi-Center Trial

Enrollment at each site exceeded the planned metrics, and it was determined that conducting a multicenter trial was feasible. On March 2, 2025 the investigators revised the trial protocol from a three-center trial focused on feasibility to a definitive multicenter randomized trial evaluating the effect of the trial interventions on clinical outcomes. Neither the investigators nor the DSMB reviewed any analyses of trial data as part of the transition from a pilot trial to a definitive multicenter trial. At the time of the transition the sample size was re-estimated to provide sufficient power to detect a difference in the revised primary outcome, the time to successful ECMO weaning, as below.

Final Sample Size Estimate for the Complete Multicenter Stage

Using simulations on data from prior patients receiving V-V ECMO in the trial setting, the investigators calculated that a total sample size of 225 patients (112 or 113 per group) would provide 80% statistical power at a two-sided alpha of 0.05 to detect a hazard ratio of 1.68 for successful ECMO weaning in the ECMO-free protocol group compared to the usual care group, using a subdistribution hazard method accounting for death as a competing event. The minimum clinically important difference in the time to successful ECMO weaning that would be required to justify routine use of the ECMO-free protocol in clinical care is unknown. The effect size of 1.68 targeted in the ECMO-Free trial is smaller (more conservative) than the effect size targeted in prior multi-center trials of V-V ECMO^1^. An effect size of 1.68 was estimated to be a difference between groups of 1.3 days in time to decannulation from ECMO.

#### **8. Effect Modification (Subgroup Analyses)**

In accordance with the Instrument for assessing the Credibility of Effect Modification Analyses (ICEMAN) recommendations^2^, we have prespecified the following variables as potential effect modifiers and hypothesized the direction of effect modification for each:

1. Age (Continuous) – if dichotomized for presentation in forest plot then we will present as age greater or equal to 50 years versus age less than 50 years. We hypothesize that age at enrollment will not modify the effect of trial group assignment on the primary outcome.
2. Body mass index (BMI) (weight [kg] / height [m]^2^) (Continuous) – if dichotomized for presentation in forest plot then we will present as BMI greater or equal to 30 versus BMI less than 30. We hypothesize that BMI will modify the effect of trial group assignment on the effect of the primary outcome, with a greater decrease in the time to successful decannulation from ECMO (in the ECMO-free protocol group compared to the usual care group) among patients with high BMI, compared to low BMI. This is based on the physiologic rationale that the severity of acute respiratory distress may be overestimated in obese patients relative to patients without obesity^3,4^, possibly leading to delayed weaning/decannulation among patients with high BMI and a greater benefit from the ECMO-free protocol among patients with high BMI.
3. Primary indication for ECMO (Acute respiratory distress syndrome / Post-lung transplant / Other). We hypothesize that primary indication for ECMO will modify the effect of trial group assignment on the primary outcome, with a greater decrease in the time to successful decannulation (in the ECMO-free protocol group compared to the usual care group) among patients with acute respiratory distress syndrome (ARDS), compared to patients with other indications. This hypothesis is supported by clinical experience and limited evidence suggesting that patients with ARDS have higher severity of disease and require longer durations of ECMO support compared to patients receiving ECMO after lung transplant^5-7^ and for other indications. This suggests that patients with ARDS may have a greater opportunity to benefit from the ECMO-free protocol.
4. Compliance at cannulation ([tidal volume/(plateau pressure-PEEP)] for patients receiving volume control; [tidal volume/driving pressure] for patients receiving pressure control) (Continuous) – if dichotomized for presentation in forest plot then we will present as compliance greater or equal to 20 mL/H_2_O versus compliance less than 20 mL/H_2_O. We hypothesize that the compliance at cannulation will modify the effect of trial group assignment on the primary outcome, with a greater decrease in the time to successful decannulation (in the ECMO-free protocol group compared to the usual care group) among patients with low compliance. This hypothesis is based on the physiologic rationale that patients with low compliance may require higher ventilator and ECMO support on average, leading to delayed assessments of readiness for decannulation from ECMO. Further, patients with low compliance may have higher severity of disease and require longer durations of ECMO support compared to patients with high compliance. This suggests that patients with low compliance may have a greater opportunity to benefit from the ECMO-free protocol.
5. Partial pressure of arterial oxygen to fraction of inspired oxygen ratio at cannulation (P:F) (Continuous) – if dichotomized for presentation in forest plot then we will present as P:F greater or equal to 60 versus P:F less than 60. We hypothesize that P:F at cannulation will modify the effect of trial group assignment on the primary outcome, with a greater decrease in the time to successful decannulation (in the ECMO-free protocol group compared to the usual care group) among patients with a lower P:F at cannulation. This hypothesis is based on the physiologic rationale that patients with a lower P:F at cannulation may require higher ventilator and ECMO support on average, leading to delayed assessments of readiness for decannulation from ECMO. Further, patients with lower P:F at cannulation may have higher severity of disease and require longer durations of ECMO support compared to patients with high P:F. This suggests that patients with low P:F may have a greater opportunity to benefit from the ECMO-free protocol.
6. pH at cannulation (Continuous) – if dichotomized for presentation in forest plot then we will present as pH greater or equal to 7.25 versus pH less than 7.25. We hypothesize that pH at cannulation will modify the effect of trial group assignment on the primary outcome, with a greater decrease in the time to successful decannulation (in the ECMO-free protocol group compared to the usual group) among patients with a low pH at cannulation. This hypothesis is based on the physiologic rationale that patients with lower pH at cannulation may require higher ventilator and ECMO support, leading to delayed assessments of readiness for decannulation from ECMO. Further, patients with lower pH at cannulation may have higher severity of disease and require longer durations of ECMO support compared to patients with higher pH. This suggests that patients with low pH may have a greater opportunity to benefit from the ECMO-free protocol.
7. Respiratory ECMO Survival Prediction (RESP) score at cannulation (Continuous) – if dichotomized for presentation in forest plot then we will present as RESP score greater or equal to 0 versus RESP score less than 0. We hypothesize that RESP score at cannulation will not modify the effect of trial group assignment on the primary outcome.

#### **9. Safety Monitoring and Adverse events**

Assuring patient safety is an essential component of the ECMO-Free Trial. An assessment of readiness for liberation from V-V ECMO is a standard-of-care intervention that has been used in clinical practice for decades with an established safety profile. However, any trial conducted for critically patients receiving a high-risk like V-V ECMO raises unique safety considerations. This trial addresses these considerations through:

1. Duplicated exclusion and daily safety screen criteria to identify and exclude patients at high risk of complications from a zero-sweep gas flow trial (e.g., those receiving arterial ECMO support).
2. Robust safety screen criteria designed to prevent performance of an ECMO-free trial for any patient in whom reduction of fraction of delivered oxygen or cessation of sweep gas flow would be unsafe.
3. Required presence of trained study personnel at bedside during initiation of the ECMO-free trial.
4. Exclusion or withdrawal of patients for whom ECMO decannulation is not intended (patients receiving ECMO as a bridge to transplant).
5. Allowance for study exclusion and withdrawal from the study based on the discretion of the patient’s treating clinicians.
6. Systematic collection of safety outcomes relevant to the study intervention.
7. Structured reporting of adverse events.

#### **10. Plan for Communication of Protocol Changes**

Any changes to the trial protocol, including changes to eligibility, outcomes, and analyses, will be implemented via a new version of the full trial protocol, tracked with the date of the update, and the version number of the trial protocol. A list summarizing the changes made with each protocol revision will be included at the end of each protocol. The updated protocol will be sent to the sIRB for approval and tracking prior to implementation of the protocol change. At the time of publication, the original trial protocol and the final trial protocol, including the summary of changes made with each protocol version, will be included in the supplementary material for publication.

#### **11. Patient Privacy, Data Storage, and Sharing**

Federal regulations 45 CFR 46 111 (a) (7) requires that, when appropriate, there are adequate provisions to protect the privacy of patients and maintain confidentiality of the data. Data will be entered directly into a secure online database. All data will be maintained in the secure online database (REDCap) until the time of study publication. No additional data are obtained beyond that which are obtained at the bedside or from the electronic medical record for the primary study. At no time during the course of this study, its analysis, or its publication will patient identities be revealed in any manner. The minimum necessary data containing patient identities are collected. All patients are assigned a unique study ID number for tracking. Data collected from the medical record is entered into REDCap. At the time of publication, a de-identified version of the database will be generated. Deidentified data will be available for sharing following trial publication with the requirements and terms of data sharing specified in the trial results manuscript.

#### **12. Trial Status**

The ECMO-Free Trial is a pragmatic, multi-center, non-blinded randomized clinical trial comparing the ECMO-free protocol vs usual care. Enrollment began on September 7, 2022, and is expected to conclude in March 2026.
